# Spatial and temporal variation in respiratory syncytial virus (RSV) subtype RNA in wastewater, and relation to clinical specimens

**DOI:** 10.1101/2024.03.16.24304392

**Authors:** Winnie Zambrana, ChunHong Huang, Daniel Solis, Malaya K. Sahoo, Benjamin A. Pinsky, Alexandria B. Boehm

**Affiliations:** Department of Civil & Environmental Engineering, Stanford University, 473 Via Ortega, Stanford, CA, USA, 94305; Department of Pathology, Stanford University School of Medicine, Stanford CA, 94305; Department of Medicine, Division of Infectious Diseases and Geographic Medicine, Stanford University School of Medicine, Stanford CA, 94305

**Keywords:** RSV A, RSV B, wastewater surveillance, subtype variability, subtype monitoring, respiratory syncytial virus

## Abstract

Respiratory syncytial virus (RSV) causes a large burden of respiratory illness, globally. It has two subtypes, RSV A and RSV B, but little is known regarding the predominance of these subtypes during different seasons and their impact on morbidity and mortality. Using molecular methods, we quantified RSV A and RSV B RNA in wastewater solids across multiple seasons and metropolitan areas to gain insight into the predominance of RSV subtypes. We determined the predominant subtype for each group using the proportion of RSV A to total RSV (RSV A + RSV B) in each wastewater sample (P_A,WW_), and conducted a comparative analysis temporally, spatially and against clinical specimens. A median P_A,WW_ of 0.00 in the first season and 0.58 in the second season indicated a temporal shift in the predominant subtype. Spatially, while we observed dominance of the same subtype, P_A,WW_ was higher in some areas (P_A,WW_ = 0.58 to 0.88). The same subtype predominated in wastewater and clinical samples, but clinical samples showed higher levels of RSV A (RSV A positivity in clinical samples = 79%, median P_A,WW_ = 0.58). These results suggest that wastewater, alongside clinical data, holds promise for enhanced subtype surveillance.

**Importance:** Respiratory syncytial virus (RSV) causes a large burden of respiratory illness, globally. It has two subtypes, RSV A and RSV B, but little is known regarding the predominance of these subtypes during different seasons and their impact on morbidity and mortality. The study illustrates that information on subtype predominance can be gleaned from wastewater. As a biological composite sample from the entire contributing population, wastewater monitoring of RSV A and B can complement clinical surveillance of RSV.

## 1. Introduction

Respiratory syncytial virus (RSV) is a leading cause of lower respiratory tract infection across all age groups. Infants, young children, and older adults are the groups with highest risk of developing severe complications from an RSV infection^1^. An RSV infection typically causes flu-like symptoms such as congestion, cough and fever, and can cause bronchiolitis and pneumonia in severe cases^2^. Each year in the United States, RSV causes between 58,000 to 80,000 children hospitalizations, and 6,000 to 10,000 older adults deaths^3,4^. It is estimated that over 100,000 deaths of children under the age of five are attributed to RSV annually globally^5^. The true burden of RSV, however, is substantially underestimated, as such an estimate relies on clinical testing of patients, often only including patients with more severe symptoms that have access to and seek medical treatment. Most older children and adults infected with RSV are excluded from these estimates, as they often only experience mild symptoms^6^. An RSV infection can also be asymptomatic which also contributes to the incomplete understanding of the true burden^7^.

RSV is an enveloped, single stranded, negative sense RNA virus that infects humans, and has two subtypes, RSV A and RSV B^8^. The major difference between subtypes is the variation in the G glycoprotein (G protein) that makes up part of the envelope^9^. The pharmaceutical interventions available to combat RSV, such as prophylactic treatments and the recently approved vaccines, consider their difference in structure and can be cross-reactive for both subtypes^10–12^. Regardless, these are only available to a limited section of the population (e.g. individuals older than 60 years old, pregnant individuals, and infants younger than 19 months)^13–17^ and have even encountered supply issues^18,19^.

Despite the known structural differences between RSV subtypes, the variability in subtype predominance remains poorly understood. The predominant subtype during an outbreak is often not characterized, as routine surveillance of RSV typically does not include subtype monitoring^20^. Studies show that although co-circulation of both subtypes is expected, typically one subtype predominates in a particular season^21^. Some epidemiological studies have found that seasons in which RSV A dominated started earlier in the winter, peaked faster, and lasted longer than seasons in which RSV B dominated^22^, while others found no difference between RSV A- a.nd RSV B-dominated seasons^23^. There is also a limited understanding of the difference in virulence between subtypes. Several epidemiological studies found that RSV A was associated with a more severe disease than RSV B, while others reported the opposite or found no difference between them^10^. Understanding how subtype predominance varies could aid in discovering potential differences in infection dynamics and virulence caused by each subtype, and could contribute to a better understanding of RSV spread.

Wastewater may represent a means for understanding the variability in RSV subtype circulation. Wastewater has recently emerged as a tool for monitoring the disease levels of a community contributing to it, as concentrations of viral genetic material in wastewater have been shown to correlate with positivity rates or clinical case rates for a number of different viruses^24–27^. Several studies have measured RSV RNA wastewater concentrations^25,28–35^, with only three of them measuring RSV subtypes using PCR-methods^25,29,30^, and one using a sequence-based methods^32^. Overall, there is limited research investigating the variability of RSV subtype predominance in wastewater. While some epidemiological studies have studied variations in subtype dominance^22,23,36,37^, their scope has been confined to clinical samples, typically encompassing individuals with severe RSV cases. This limitation is crucial when considering that the majority of individuals infected with RSV do not seek medical treatment^38^. Wastewater data bypasses biases inherently included in clinical case data and, thus, could be used to understand variations in RSV subtype predominance at a broader population scale.

The goal of this study is to investigate temporal and spatial variations in the predominant RSV subtype detected in wastewater solids samples. This investigation involves measuring concentrations of both RSV A and RSV B in wastewater solids samples across multiple seasons and metropolitan areas, determining the predominant subtype for each, and conducting a comparative analysis temporally, spatially and against clinical data.

## 2. Materials and Methods

### Study Design

This study was carried out at different wastewater treatment facilities across the United States (Table 1), comprising 240 wastewater solids samples in total. The samples in this study are part of a larger wastewater surveillance effort which measures total RSV (RSV A + RSV B) routinely in wastewater solids^39^, and were selected due to their high total RSV concentration (more information in SI). For each wastewater solids sample, we measured the concentrations of both RSV A and RSV B using digital droplet reverse transcription polymerase chain reaction (RT-PCR), and calculated the main outcome of this study — the proportion of RSV A to total RSV (P_A,WW_):

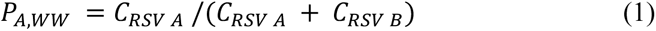

**Table 1.**
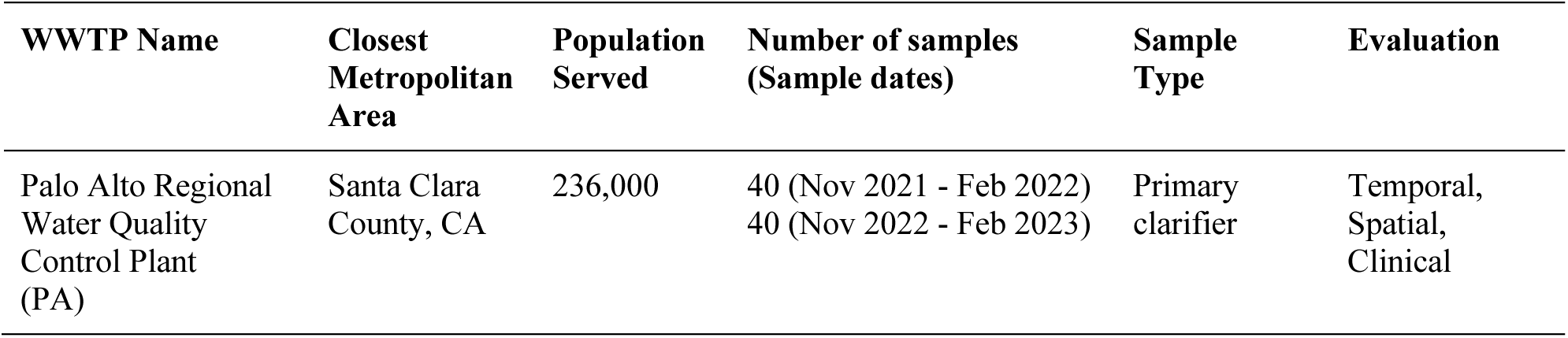
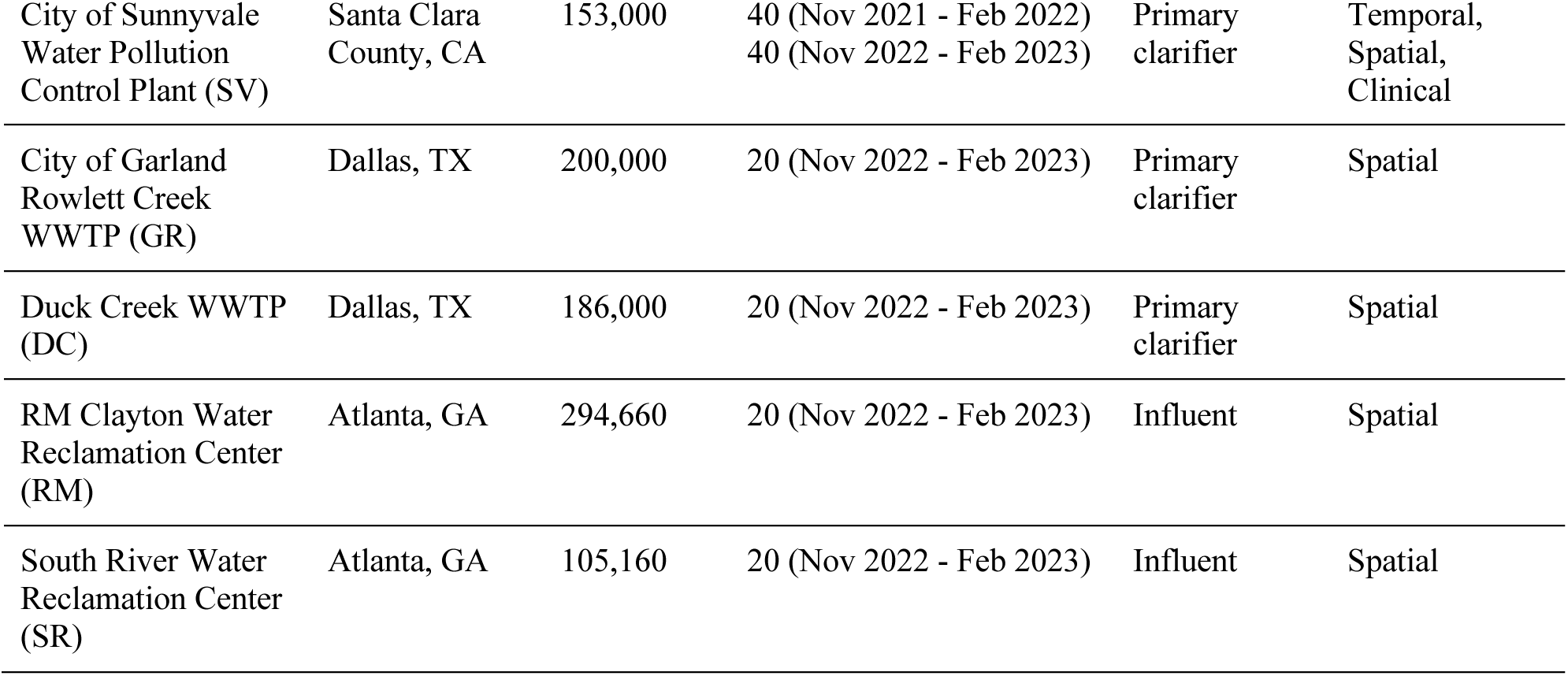
Characteristics of wastewater treatment plants (WWTPs) included in the study, number of samples per WWTP, sample type, and the evaluations in which they were considered. Settled solids were collected directly from the primary clarifier (“primary clarifier”), or were collected from a raw influent sample (“influent”) and allowed to settle for 10-15 minutes in the laboratory, and later aspirated into a falcon tube using a serological pipette.

| WWTP Name | Closest Metropolitan Area | Population Served | Number of samples (Sample dates) | Sample Type | Evaluation |
| --- | --- | --- | --- | --- | --- |
| Palo Alto Regional Water Quality Control Plant (PA) | Santa Clara County, CA | 236,000 | 40 (Nov 2021 - Feb 2022)<br>40 (Nov 2022 - Feb 2023) | Primary clarifier | Temporal, Spatial, Clinical |
| City of Sunnyvale<br>Water Pollution<br>Control Plant (SV) | Santa Clara<br>County, CA | 153,000 | 40 (Nov 2021 - Feb 2022)<br>40 (Nov 2022 - Feb 2023) | Primary<br>clarifier | Temporal,<br>Spatial,<br>Clinical |
| City of Garland<br>Rowlett Creek<br>WWTP (GR) | Dallas, TX | 200,000 | 20 (Nov 2022 - Feb 2023) | Primary<br>clarifier | Spatial |
| Duck Creek WWTP<br>(DC) | Dallas, TX | 186,000 | 20 (Nov 2022 - Feb 2023) | Primary<br>clarifier | Spatial |
| RM Clayton Water<br>Reclamation Center<br>(RM) | Atlanta, GA | 294,660 | 20 (Nov 2022 - Feb 2023) | Influent | Spatial |
| South River Water<br>Reclamation Center<br>(SR) | Atlanta, GA | 105,160 | 20 (Nov 2022 - Feb 2023) | Influent | Spatial |

where *C_RSV A_* is the concentration of RSV A, and *C_RSV B_* is the concentration of RSV B in the sample in units of copies/g dry weight. A proportion greater than 0.5, for example, indicates that RSV A is the predominant subtype for that sample. Equation 1 assumes that *C_RSV Total_* = *C_RSV A_* + *C_RSV B_* in each sample. Shedding of RSV A and RSV B by infected individuals into the wastewater stream is assumed to be similar due to the lack of information in the literature^40^.

#### Temporal Evaluation

We compared P_A,WW_ between two different RSV seasons (2021-2022 and 2022-2023) using wastewater solids samples collected from two WWTPs in the Santa Clara County, CA area (PA and SV). Samples from Season 1 (2021-2022) were collected between November 15, 2021 and February 28, 2022, and samples from Season 2 (2022-2023) were collected between November 1, 2022, and February 28, 2023 (Table 1). The WWTPs included in this evaluation are located 20 kilometers (km) from each other and have neighboring sewersheds (Figure 1). In a secondary evaluation, we compared results between these WWTPs within each season to determine if samples from the WWTPs, located in close proximity (∼20 km apart with neighboring sewersheds), yield similar results.

**Figure 1.**
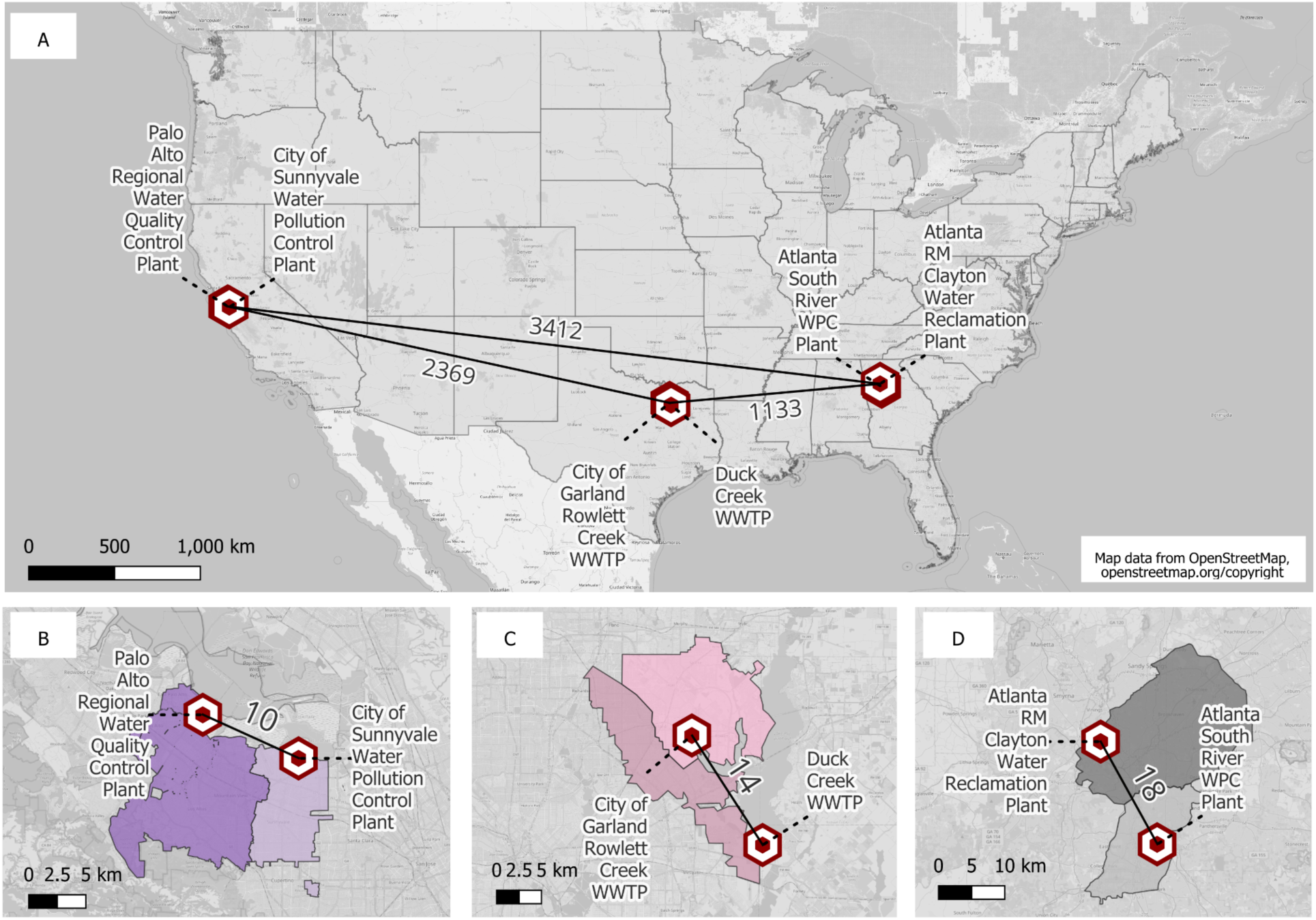
Wastewater treatment plants included in this study. Distance (km) between each of the three metropolitan areas evaluated **(A)**. Distance (km) between both WWTPs selected to represent each area and their corresponding sewersheds: Santa Clara County, California **(B)**, Dallas, Texas **(C)**, and Atlanta, Georgia **(D)**. WWTPs shown in Figure 1B are part of all three evaluations, and WWTPs shown in Figures 1C-1D are only included in the spatial evaluation. This figure was generated using QGIS; map layer from OpenStreetMap (openstreetmap/org/copyright).

#### Spatial Evaluation

We compared P_A,WW_ between WWTPs located across the United States using wastewater solids samples collected during a single RSV season (2022-2023). Wastewater solids samples were collected between November 1, 2022 and February 28, 2023 from three United States areas: Santa Clara County, CA, Dallas, TX, and Atlanta, GA (Table 1). These areas were selected as they are considered some of the most populous metropolitan cities in the United States^41^, and each have two WWTPs with neighboring sewersheds. Each metropolitan area was represented by two different WWTPs located within 20 km from each other and with neighboring sewersheds. The distances between each metropolitan area were approximately 1,000 km, 2,000 km, and 3,000 km (Figure 1). Note that the samples from Santa Clara County, CA used for this analysis were the same ones used for the temporal variation analysis described above.

#### Clinical Evaluation

We compared the relative proportion of RSV A identified using wastewater solids (P_A,WW_) to the fraction of clinical specimens collected from individuals residing within and adjacent to the sewersheds in Santa Clara County, CA identified as subtype A (F_A, Cl_). A total of 80 wastewater solids samples and 602 clinical samples were included in this evaluation (Table 1), and all were collected in the 2022-2023 RSV season. The wastewater solids samples were already described as those used in the temporal evaluation study. Clinical samples, collected between October 1, and December 31, 2022, were processed by the Stanford Clinical Virology Laboratory (Palo Alto, CA). The laboratory receives samples from patients in the Santa Clara, CA area, including patients living and/or working within the sewershed of the selected WWTPs. This study was conducted with Stanford Institutional Review Board approval (protocol 62834), and individual consent was waived.

Using the total number of clinical samples subtyped, we calculated the fraction of clinical specimens identified as subtype A to the total number identified as RSV positive (F_A, Cl_):

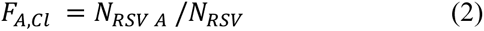

Where *N_RSV A_* is the number of samples that were positive for RSV A, and *N_RSV_* is the total number of samples that were subtyped and found positive for either RSV A or RSV B, *N_RSV_* = *N_RSV A_* + *N_RSV b_*. F_A, Cl_ was compared to P_A,WW_.

#### Wastewater solids storage and freeze-thaw cycles impact assessment

Concentrations of pepper mild mottle virus (PMMoV) and total RSV were measured in all wastewater samples immediately after they were collected with no storage (Table S1). To assess the impact of storage and freeze-thaw cycles in our samples, a subset of randomly selected samples (n = 30) from Sunnyvale (SV) and Palo Alto (PA) WWTPs were measured a second time for PMMoV after two freeze-thaw cycles. For all samples (n = 240), the sum of RSV A and RSV B concentrations taken after two freeze-thaw cycles were compared against the measurements of total RSV in fresh samples.

### Statistical Analysis

The median P_A,WW_ of wastewater solids samples were used to compare the predominant subtype between different groups in the statistical analyses. Non-parametric methods were employed as the wastewater data were found to be not normally distributed (Shapiro-Wilk test, W=0.90, p=9.1 x10^-12^). A Kruskal-Wallis test was used to test the null hypothesis that the P_A,WW_ was not different between groups, and a post-hoc test (Conover-Iman test) was used to compare the P_A,WW_ between groups.

A bootstrap method was implemented to compare the F_A, Cl_ to the P_A,WW_. A synthetic dataset consisting of zeros and ones was generated using the total number of clinical samples subtyped (*N*_*RSV*_) and the total number of samples identified as subtype A (*N*_*RSV A*_). Next, a collection of 1000 F_A, Cl_ was established by randomly selecting, with replacement, from the synthetic list and calculating it using equation 2. From the collection of 1000 F_A, Cl_, a subset of 80 were randomly selected (equivalent to the number of wastewater solids samples), and subsequently used in the Kruskal-Wallis and post-hoc analyses^43^.

Lastly, three supplementary statistical analyses were performed. Two Kruskal-Wallis tests were conducted to compare fresh sample measurements of RSV and pepper mild mottle virus (PMMoV) with measurements from samples subjected to storage and freeze-thaw cycles to assess the impact of storage on measurements in the wastewater samples. Additionally, Kendall’s τ was used to test the null hypothesis that total RSV (RSV A +RSV B) concentration levels and P_A,WW_ are statistically independent.

Two hypotheses were tested (two tests: Kruskal-Wallis test and post-hoc test) for each comparison (temporal, spatial or clinical)] and three hypotheses were tested for the supplementary analysis, for a total of 9 hypothesis tests. A p value of 0.006 (0.05/9) for alpha = 0.05 was used to adjust for multiple comparisons (Bonferroni correction). Statistical analysis was completed using R version 4.3.0 within RStudio version 2021.09.1. Wastewater solids data from this study is available at the Stanford Digital Repository^44^.

### Procedures

#### Wastewater solids sample collection and pre-analytical processing

Wastewater solids were either grab samples of settled solids from the primary clarifier, or 24-hr composite raw influent samples in which solids were allowed to settle for 10-15 minutes in the laboratory, and aspirated into a falcon tube using a serological pipette (Table 1). Samples were collected in sterile bottles by WWTP staff, immediately stored at 4°C, delivered to the laboratory, and processed within 6 hours upon arrival. Samples were dewatered by centrifugation and resuspended in DNA/RNA shield (Zymo Research, Irvine, CA) spiked with bovine coronavirus vaccine (BCoV, Zoetis, Calf-Guard Cattle Vaccine) to a final concentration of 75 mg/ml to minimize inhibition of measurements in solids^45^. Resuspended solids were homogenized and centrifuged, and the supernatant was withdrawn for nucleic-acid extraction. A 0.5 g to 1 g aliquot of the dewatered solids was oven dried to measure its dry weight for its use in the dimensional analysis. Detailed description of the pre-analytical processing can be found elsewhere^46,47^ including in detailed protocols on protocols.io^47^.

#### Wastewater solids RNA extraction and quantification

RNA extraction from the supernatant was performed using the Chemagic Viral DNA/RNA 300 kit H96 for the Perkin Elmer Chemagic 360 (Perkin Elmer, Waltham, MA). Extract was further purified using the Zymo One-Step PCR inhibitor removal columns (Zymo Research, Irvine, CA). Nuclease-free water served as a negative extraction control, and BCoV served as a process control. RNA extracts were immediately processed (no storage or freeze-thaw) for quantification of BCoV, pepper mild mottle virus (PMMoV), and total RSV (RSV A + RSV B).

Additional aliquots of RNA extracts were stored at -80°C (for 0-700 days) and later subjected to two freeze-thaw cycles for RSV A and RSV B measurements. Although PMMoV was already measured using fresh samples, they were measured again on a subset of the stored and freeze thawed samples to assess the impact of storage and freeze-thaw cycles in our samples (as described in the Study Design section).

RNA extracts were used as template neat using one-step RT-ddPCR Advanced Kit for Probes. RSV A and RSV B were measured in a duplex assay using methods described below, and PMMoV, BCoV and total RSV were measured in fresh samples using methods described in detail in other publications^39^.

Primers and probes were purchased from Integrated DNA Technologies (IDT, Coralville, IA). Primers and probe sequences and corresponding thermal cycling conditions were selected from published assays (Table S1)^25,48–50^. The fluorescence signal was analyzed in the HEX channel for RSV A and the FAM channel for RSV B (Table S1). Nuclease-free water served as a negative PCR control, and gene blocks (Integrated DNA Technologies (IDT), Coralville, IA) served as positive controls for all targets. Non-infectious intact RSV A virus (NATRSVA-STQ, Zeptomatrix, Buffalo, NY) and non-infectious intact RSV B virus (NATRSVB-STQ, Zeptomatrix, Buffalo, NY) were used as additional positive controls for RSV A and RSV B, correspondingly. RNA target concentrations were measured via digital droplet RT-PCR (RT-ddPCR) using an Automated Droplet Generator (Bio-Rad, Hercules, CA), C1000 Touch (Bio-rad) thermocycler, and a QX200 Droplet Reader (Bio-Rad). Droplets were analyzed using QuantaSoft and QuantaSoft Analysis Pro software. Each sample had three replicate wells. Replicate wells were merged, and were required to have at least 10,000 droplets post-merger. A sample was required to have three or more positive droplets across three merged wells to be scored as positive for a target. The lower detection limit for the RSV A and RSV B assays was approximately 2,200 gc/g dry weight, assuming three positive droplets across merged wells. A detailed description of RNA extraction of resuspended solids, quantification using digital RT-ddPCR, and dimensional analysis used to convert concentrations per reaction to copies per gram of dry weight is described in detail in other publications^46,51^ and on protocols.io^52,53^.

#### Clinical samples RNA extraction and quantification

RSV-positive respiratory samples, including nasopharyngeal, mid-turbinate, and anterior nasal swabs in viral transport media, as well as bronchoalveolar lavage fluid, were extracted on the PerkinElmer Chemagic 360 instrument using the Chemagic viral DNA/RNA 300 Kit H96 according to the manufacturer’s recommendations. Samples were extracted from 300 µL and eluted in 60 µL. Samples were identified as positive for either RSV A or RSV B using reverse-transcription qPCR (RT-qPCR). The primer and probe sequences were adapted from the assay described by Wang *et al.* 2019^54^, and modified to include RNAse P primers and probe to serve as an internal control (Table S2).

RT-qPCR was performed using Invitrogen Superscript III Platinum One-Step qRT-PCR kit (Invitrogen, Carlsbad, CA) on the Bio-rad CFX-96 instrument (Bio-rad, Hercules, CA). Each 25 µL reaction contained 12.5 µL of 2X Buffer, 0.5 µL of enzyme mix, 2 µL of primer/probe mix, and 10 µL of eluate. Cycling conditions were as follows: hold at 52°C for 15 min, 94°C for 2 min, then 45 cycles of 94°C for 15 sec, 55°C for 40 sec, and 68°C for 20 sec. The fluorescence signal was analyzed in the FAM (A), Cy5 (B), and HEX (internal control: RNase P) channels. Thresholds were set at 3000, 1000, and 200 relative fluorescence units (RFU), respectively. Any exponential amplification curve crossing either the FAM or Cy5 RFU thresholds were interpreted as positive for the corresponding target. Samples were considered to have failed extraction or contain inhibitory substances if RNase P did not amplify at a cycle threshold value ≤ 35 cycles. Samples were considered to be untypable if both RSV A and RSV B were not detected.

## 3. Results and Discussion

### QA/QC

Results are reported as suggested by the Environmental Microbiology Minimal Information (EMMI) guidelines (Figure S3-S4)^55^. Negative and positive PCR and extraction controls yielded negative and positive results, respectively. Median BCoV recovery in fresh samples was around 100% (median = 110%, IQR = 69%), and PMMoV was stable across fresh samples (median = 1.16 x10^9^ gc/g, IQR = 1.19 x10^9^ gc/g).

A comparison of total RSV and PMMoV concentrations reported in this study to concentrations measured in samples with no storage and no freeze-thaw cycles showed an effect on target quantification. The median ratios of RSV and PMMoV in stored to fresh samples were 1.29 (IQR = 0.62, n = 240) and 0.40 (IQR = 0.41, n = 30), respectively. Although the ratios show less than an order of magnitude difference between measurements in fresh and stored samples, we found that fresh sample measurements for both RSV and PMMoV were significantly different from measurements from those measurements in stored samples (Kruskal-Wallis test, RSV comparison p=1.60 x 10^-3^; PMMoV comparison p= 1.69 x 10^-10^). This suggests that storage and freeze thaw might have had an impact on target measurements.

We found no significant correlation between concentrations of total RSV and P_A,WW_ (Kendall’s τ test, τ = 0.09, z = 2.05, p=0.04). This suggests that the levels of total RSV RNA detected in wastewater are not reflective of the predominance of a specific subtype.

### Overall Wastewater and Clinical Samples Results

All wastewater solids samples were positive for RSV A and/or RSV B, with the exception of three samples that were non-detects for either RSV A or RSV B. Median RSV A concentration across all wastewater samples (n = 240) was 1.31 x10^4^ gc/g dry weight (IQR = 4.07 x10^4^ gc/g), and median RSV B concentration was 1.90 x10^4^ gc/g dry weight (IQR = 2.54 x10^4^ gc/g) (Figure S1). Overall median P_A,WW_ across all seasons and all wastewater treatment plants was 0.47 (IQR = 0.74, n = 237) (Figure S2). A total of 593 of 602 RSV-positive clinical samples were identified as either RSV A or RSV B, and 9 samples were considered untypable. We identified the predominant RSV subtype across multiple wastewater treatment plants and conducted a comparative analysis temporally, spatially, and against clinical specimens.

### Temporal Evaluation

The median P_A,WW_ was significantly larger in Season 2 (2022-2023) than Season 1 (2021-2022) for both wastewater treatment plants (Palo Alto Regional Water Quality Control Plant [PA] and City of Sunnyvale Water Pollution Control Plant [SV]) (Conover-Iman test, exact p-values shown in Table S3-S4). The combined median P_A,WW_ from both WWTPs for Season 1 was 0 (PA median = 0.0, IQR = 0.11, n = 40; SV median = 0.0, IQR = 0.16, n = 40), while combined median P_A,WW_ for Season 2 was 0.58 (PA median = 0.53, IQR = 0.21, n = 40; SV median = 0.61, IQR = 0.21, n = 40) (Figure 2, Table S3-S4). The results confirm a change in the predominant subtype between two consecutive RSV seasons. RSV B was the predominant subtype during 2021-2022 with limited RSV A circulation, while in the following season (2022-2023), RSV A dominated with RSV B also circulating.

**Figure 2.**
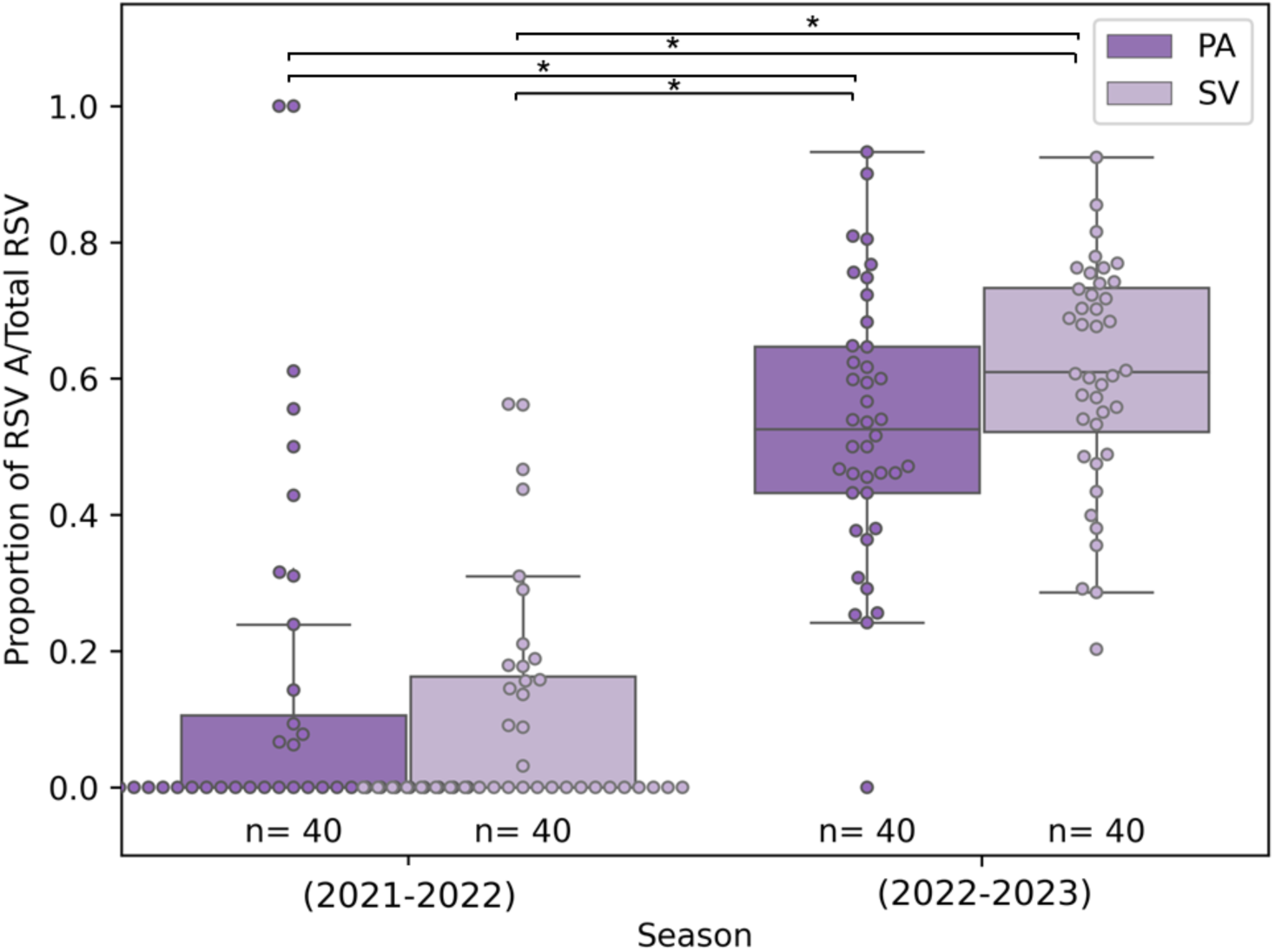
Proportion of RSV A to total RSV (P_A,WW_) for the temporal evaluation comparing Palo Alto Regional Water Quality Control Plant (PA) and City of Sunnyvale Water Pollution Control Plant (SV) for Season 1 (2021-2022) and Season 2 (2022-2023). Each box plot is made up of the 25th quartile, median, and 75th quartile proportion of RSV A total RSV for each WWTP per season, and length of each whisker 1.5 times the interquartile range(IQR). Boxplots are overlaid with jittered data points from each group.*= Statistically Significant per post-hoc (Conover−Iman) test with a significance level p= 0.006 accounting for the Bonferroni Correction.

Our results align with epidemiological studies that have also reported shifts in subtype predominance between seasons^23,36,56^. Observing these results in wastewater solids, which capture contributions from individuals with a wider spectrum of disease severity than clinical data, suggests that changes in subtype dominance between seasons are also observable at the population level. Tracking subtype predominance in wastewater each season could enhance our understanding of RSV dynamics at the population level, particularly when subtype monitoring is not integrated into routine clinical surveillance^20^.

Wastewater treatment plants with neighboring sewersheds (PA and SV) showed similar subtype patterns. For both seasons studied, we found no significant difference in the median P_A,WW_ between each wastewater treatment plant (Conover-Iman test, exact p-values shown in Table S3-S4). This result suggests that if resources are constrained, monitoring one geographic area may provide insights into subtype predominance dynamics in adjacent areas.

### Spatial Evaluation

Across the three metropolitan areas evaluated during the 2022-2023 season, RSV subtype A predominated (Santa Clara County, CA median = 0.58, IQR = 0.26, n = 80; Dallas, TX median = 0.88, IQR = 0.34, n = 39; Atlanta, GA median = 0.68, IQR = 0.74, n = 38) (Figure 3). However, the levels at which RSV A was found varied. We found that P_A,WW_ was significantly higher in Dallas, TX than in Santa Clara County, CA and Atlanta, GA, but was similar between Santa Clara County, CA and Atlanta, GA (Conover-Iman test, exact p-values shown in Table S3-S4).

**Figure 3.**
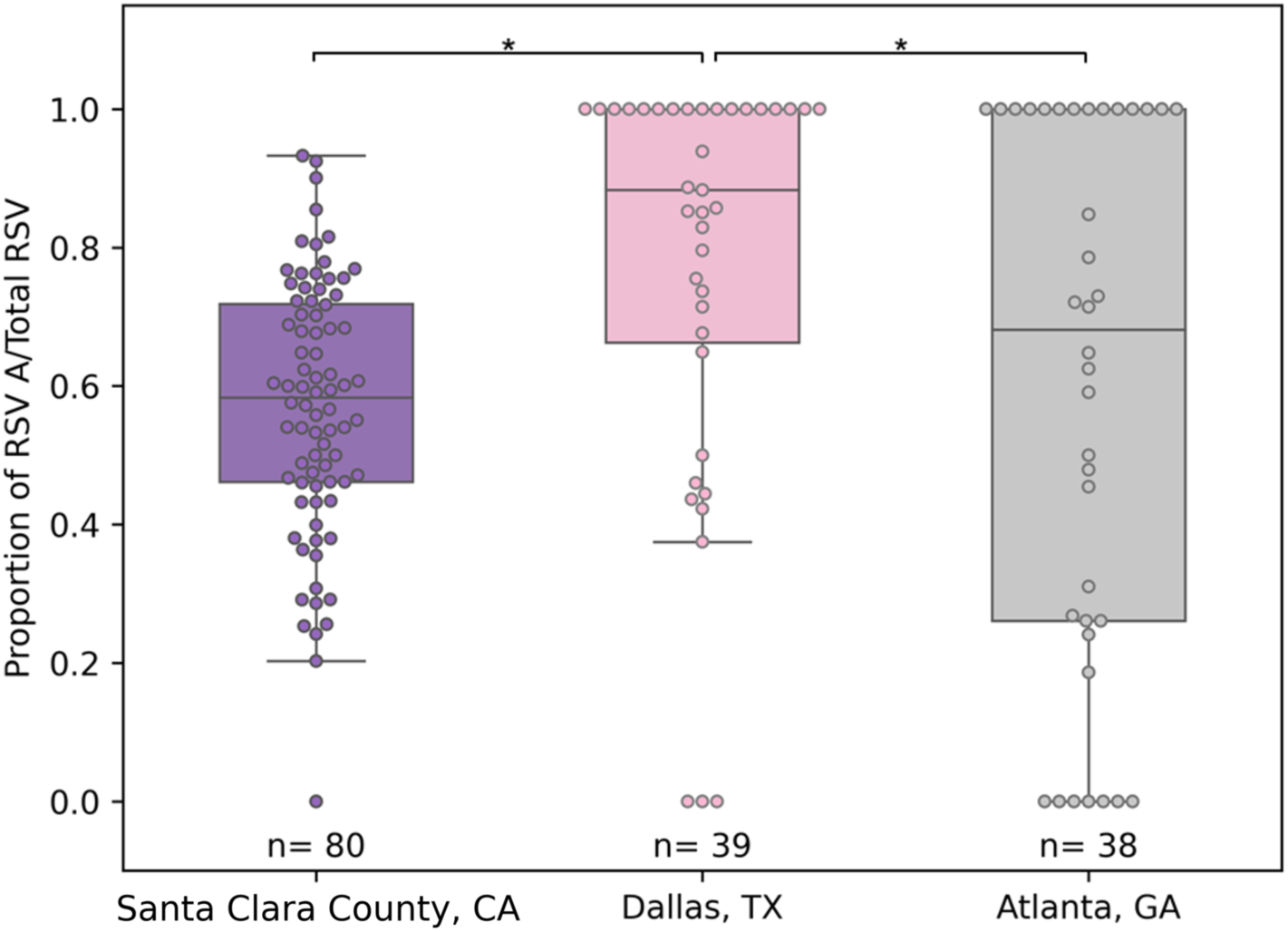
Proportion of RSV A to total RSV (P_A,WW_) for the spatial evaluation comparing wastewater solids samples from Santa Clara County, CA, Dallas, TX, and Atlanta for Season 2 (2022-2023). Each box plot is made up of the 25th quartile, median, and 75th quartile proportion of RSV A total RSV for two WWTPs per area, and length of each whisker 1.5 times the interquartile range(IQR). Boxplots are overlaid with jittered data points from each group.*= Statistically Significant per post-hoc (Conover−Iman) test with a significance level p= 0.006 accounting for the Bonferroni Correction.

Our results, showing dominance of the same RSV subtype across the three metropolitan areas, are consistent with epidemiological studies using clinical samples that have found geographic clustering of viral lineages in large geographical areas (e.g., Australian states)^36^, and that have concluded that variants are restricted to a local level (i.e., country-wide rather than global)^37^.

Despite sharing a predominant subtype, we observed a lack of association between geographic proximity and levels of one specific subtype. The areas closest to each other (∼1,000 km and ∼2,000 km apart) were found to have different P_A,WW_, and the two areas with furthest distance from each other (∼3,000 km apart) were found to follow similar subtype patterns. Factors beyond physical distance — such as environmental, demographic and transit accessibility factors — may play a crucial role in shaping the levels of subtype dominance. For instance, local climate, minimum temperature, precipitation, and population density have been shown to influence RSV dynamics including onset and timing of peak infectivity^57,58^, and urban public transportation systems have been shown to facilitate transmission of respiratory illnesses^59^. The three metropolitan areas included in this study experience similar temperate climates with mild winters, but experience different levels of precipitation^60,61^, have different population densities^62^, and have varying levels of public transportation usage^63^ which could have contributed to the difference in P_A,WW_.

### Clinical Evaluation

During the 2022-2023 season, 79% (470/593) of the clinical RSV-positive samples were subtyped as RSV A (bootstrapping F_A, Cl_ median = 0.79, IQR = 0.03, n =80). Wastewater solids samples had a median P_A,WW_ of 0.58 (IQR = 0.26, n = 80). Therefore, subtype A dominated in both sample types. However, we found the levels of subtype A in clinical samples was significantly higher than in wastewater solids samples (Conover-Iman test, p=3.88 x 10^-26^) (Table S3-S4).

To our knowledge, this is the first study to confirm that wastewater data reflects the same RSV subtype dominance as clinical data. Our results highlight the validity of wastewater as an appropriate epidemiological tool to monitor subtype predominance within a community. The discrepancies in specific subtype levels observed between clinical and wastewater solids samples may stem from the broader representation in wastewater, encompassing mild and asymptomatic cases. Clinical samples, in contrast, might have a larger representation of pediatric infections when considering that children under two are less likely contributors to wastewater solids samples. Wastewater data, in combination with clinical data, could help to provide a comprehensive understanding of the RSV burden.

### Limitations and Future Work

This study has several limitations. First, this study was conducted during atypical RSV periods due to the COVID-19 pandemic and the implementation of nonpharmaceutical interventions. While both seasons began earlier in the year than prepandemic levels, the 2021-2022 season lasted longer, and the 2022-2023 season exhibited a higher peak number of cases compared to typical RSV epidemics^64^. Another limitation is that we were not able to account for any potential impact of the newly approved vaccines or prophylactic antibody on RSV dynamics, as they were approved for public usage in certain populations subsequent to the 2022-2023 season^14,16^. Additionally, the temporal evaluation was conducted using data from only two RSV seasons, and the spatial evaluation was limited to three metropolitan areas. A wider range of seasons across a more extensive array of metropolitan areas would enhance our conclusions regarding spatial-temporal subtype predominance. Due to the nature of this retrospective study, another limitation is that wastewater solids samples used to detect RSV subtypes were subjected to storage and freeze thaw which might have had an impact on target measurements. An additional limitation is that the clinical data might also include individuals residing outside the sewershed from the selected WWTPs which could potentially bias the clinical evaluation results. Lastly, although wastewater captures a larger percent of the population infected with RSV than clinical data, it is likely that children under the age of two contribute minimally to wastewater solids samples; therefore, wastewater provides a comprehensive yet incomplete picture of the burden.

Despite the aforementioned limitations, the findings in this study suggest that future work regarding RSV subtype variability in several areas is warranted. Future studies could conduct finer-scale temporal evaluations of subtype predominance variability in wastewater, examining potential differences in onset and peak timings between seasons dominated by different subtypes. Spatial evaluations could consider differences in environmental and/or demographic conditions between areas studied that might influence subtype predominance such as climate, precipitation, population density, public transportation usage and other factors. Investigations comparing subtype predominance in wastewater to clinical data could include patient characteristics, distinguishing between pediatric and non-pediatric cases, as well as in-patient vs outpatient cases, to determine the different factors that affect subtype dominance in each signal. Lastly, wastewater monitoring could be integrated into long-term epidemiological studies investigating variability, infection dynamics, virulence characteristics of RSV subtypes, or be used to investigate the effectiveness of the newly approved vaccines at the population level. Wastewater could provide a vital comprehensive picture of cases ranging in severity, and combined with clinical data, could be robust tools to better understand these complex issues.

## 4. Conclusions

This study presents an analysis of RSV subtypes in wastewater across multiple seasons, metropolitan areas, and in comparison to clinical specimens. We found that the predominant RSV subtype varied temporally, but remained consistent spatially and between wastewater and clinical samples. Temporal evaluation revealed a shift in the predominant subtype between consecutive seasons. Spatially, while we observed dominance of the same subtype across metropolitan areas, the levels of one specific subtype varied, indicating that factors beyond geographic proximity may influence levels of subtype prevalence. When comparing wastewater samples to clinical samples, we found that the same subtype dominated in both sample types, but presence of a specific subtype was higher in clinical samples. Our findings suggest that wastewater, in conjunction with clinical data, holds promise for enhanced subtype surveillance, understanding subtype dynamics, and controlling RSV spread. Future research in RSV subtype predominance can investigate finer-scale temporal variations, study characteristics beyond physical distance, incorporate patient characteristics into clinical-wastewater comparisons, and implement wastewater monitoring into epidemiological studies.

## Data Availability

All wastewater data are available online at the Stanford Digital Repository. Clinical data are available upon request to the authors.

https://doi.org/10.25740/jb062yw6825

## 5. Supporting Material

Additional details about the methods and results used in this study (Tables S1-S4 and Figures S1-S4).

## 6. Acknowledgements

We acknowledge the wastewater treatment plants that kindly provided samples for this study.

## Notes

### Competing Interest Statement

The authors have declared no competing interest.

### Funding Statement

This study was funded by a Stanford Graduate Fellowship and the Sergey Brin Family Foundation.

### Author Declarations

IRB of Stanford University gave ethical approval for this work; IRB 62834.

## 7. References

(1) Respiratory Syncytial Virus (RSV) disease. World Health Organization. https://www.who.int/teams/health-product-policy-and-standards/standards-and-specifications/vaccine-standardization/respiratory-syncytial-virus-disease (accessed 2023-07-25).

(2) Symptoms and Care for RSV. Centers for Disease Control and Prevention. https://www.cdc.gov/rsv/about/symptoms.html (accessed 2023-07-25).

(3) Learn about RSV in Infants and Young Children. Centers for Disease Control and Prevention. https://www.cdc.gov/rsv/high-risk/infants-young-children.html (accessed 2023-07-25).

(4) Learn about RSV in older adults with chronic medical conditions. Centers for Disease Control and Prevention. https://www.cdc.gov/rsv/high-risk/older-adults.html (accessed 2023-07-25).

(5) Li, Y.; Wang, X.; Blau, D. M.; Caballero, M. T.; Feikin, D. R.; Gill, C. J.; Madhi, S. A.; Omer, S. B.; Simões, E. A. F.; Campbell, H.; Pariente, A. B.; Bardach, D.; Bassat, Q.; Casalegno, J.-S.; Chakhunashvili, G.; Crawford, N.; Danilenko, D.; Do, L. A. H.; Echavarria, M.; Gentile, A.; Gordon, A.; Heikkinen, T.; Huang, Q. S.; Jullien, S.; Krishnan, A.; Lopez, E. L.; Markić, J.; Mira-Iglesias, A.; Moore, H. C.; Moyes, J.; Mwananyanda, L.; Nokes, D. J.; Noordeen, F.; Obodai, E.; Palani, N.; Romero, C.; Salimi, V.; Satav, A.; Seo, E.; Shchomak, Z.; Singleton, R.; Stolyarov, K.; Stoszek, S. K.; Gottberg, A. von; Wurzel, D.; Yoshida, L.-M.; Yung, C. F.; Zar, H. J.; Abram, M.; Aerssens, J.; Alafaci, A.; Balmaseda, A.; Bandeira, T.; Barr, I.; Batinović, E.; Beutels, P.; Bhiman, J.; Blyth, C. C.; Bont, L.; Bressler, S. S.; Cohen, C.; Cohen, R.; Costa, A.-M.; Crow, R.; Daley, A.; Dang, D.-A.; Demont, C.; Desnoyers, C.; Díez-Domingo, J.; Divarathna, M.; Plessis, M. du; Edgoose, M.; Ferolla, F. M.; Fischer, T. K.; Gebremedhin, A.; Giaquinto, C.; Gillet, Y.; Hernandez, R.; Horvat, C.; Javouhey, E.; Karseladze, I.; Kubale, J.; Kumar, R.; Lina, B.; Lucion, F.; MacGinty, R.; Martinon-Torres, F.; McMinn, A.; Meijer, A.; Milić, P.; Morel, A.; Mulholland, K.; Mungun, T.; Murunga, N.; Newbern, C.; Nicol, M. P.; Odoom, J. K.; Openshaw, P.; Ploin, D.; Polack, F. P.; Pollard, A. J.; Prasad, N.; Puig-Barberà, J.; Reiche, J.; Reyes, N.; Rizkalla, B.; Satao, S.; Shi, T.; Sistla, S.; Snape, M.; Song, Y.; Soto, G.; Tavakoli, F.; Toizumi, M.; Tsedenbal, N.; Berge, M. van den; Vernhes, C.; Mollendorf, C. von; Walaza, S.; Walker, G.; Nair, H. Global, Regional, and National Disease Burden Estimates of Acute Lower Respiratory Infections Due to Respiratory Syncytial Virus in Children Younger than 5 Years in 2019: A Systematic Analysis. The Lancet 2022, 399 (10340), 2047–2064. 10.1016/S0140-6736(22)00478-0.

(6) Tin Tin Htar, M.; Yerramalla, M. S.; Moïsi, J. C.; Swerdlow, D. L. The Burden of Respiratory Syncytial Virus in Adults: A Systematic Review and Meta-Analysis. Epidemiol. Infect. 148, e48. 10.1017/S0950268820000400.

(7) Munywoki, P. K.; Koech, D. C.; Agoti, C. N.; Bett, A.; Cane, P. A.; Medley, G. F.; Nokes, D. J. Frequent Asymptomatic Respiratory Syncytial Virus Infections During an Epidemic in a Rural Kenyan Household Cohort. J. Infect. Dis. 2015, 212 (11), 1711–1718. 10.1093/infdis/jiv263.

(8) Carvajal, J. J.; Avellaneda, A. M.; Salazar-Ardiles, C.; Maya, J. E.; Kalergis, A. M.; Lay, M. K. Host Components Contributing to Respiratory Syncytial Virus Pathogenesis. Front. Immunol. 2019, 10, 2152. 10.3389/fimmu.2019.02152.

(9) Yu, J.-M.; Fu, Y.-H.; Peng, X.-L.; Zheng, Y.-P.; He, J.-S. Genetic Diversity and Molecular Evolution of Human Respiratory Syncytial Virus A and B. Sci. Rep. 2021, 11 (1), 12941. 10.1038/s41598-021-92435-1.

(10) Vandini, S.; Biagi, C.; Lanari, M. Respiratory Syncytial Virus: The Influence of Serotype and Genotype Variability on Clinical Course of Infection. Int. J. Mol. Sci. 2017, 18 (8), 1717. 10.3390/ijms18081717.

(11) Hashimoto, K.; Hosoya, M. Neutralizing Epitopes of RSV and Palivizumab Resistance in Japan. Fukushima J. Med. Sci. 2017, 63 (3), 127–134. 10.5387/fms.2017-09.

(12) Che, Y.; Gribenko, A. V.; Song, X.; Handke, L. D.; Efferen, K. S.; Tompkins, K.; Kodali, S.; Nunez, L.; Prasad, A. K.; Phelan, L. M.; Ammirati, M.; Yu, X.; Lees, J. A.; Chen, W.; Martinez, L.; Roopchand, V.; Han, S.; Qiu, X.; DeVincenzo, J. P.; Jansen, K. U.; Dormitzer, P. R.; Swanson, K. A. Rational Design of a Highly Immunogenic Prefusion-Stabilized F Glycoprotein Antigen for a Respiratory Syncytial Virus Vaccine. Sci. Transl. Med. 2023, 15 (693), eade6422. 10.1126/scitranslmed.ade6422.

(13) RSV information for healthcare providers. Centers for Disease Control and Prevention. https://www.cdc.gov/rsv/clinical/index.html (accessed 2023-07-10).

(14) Commissioner, O. of the. FDA Approves First Respiratory Syncytial Virus (RSV) Vaccine. FDA. https://www.fda.gov/news-events/press-announcements/fda-approves-first-respiratory-syncytial-virus-rsv-vaccine (accessed 2023-06-29).

(15) Regan, A. That Respiratory Infection You Had That Wasn’t COVID Has Never Had a Vaccine—until Now. Fortune Well. https://fortune.com/well/2023/05/10/is-there-an-rsv-vaccine-arexvy-gsk-fda-approval/ (accessed 2023-06-29).

(16) Jewett, C. F.D.A. Approves Pfizer’s R.S.V. Vaccine for Older Adults. The New York Times. May 31, 2023. https://www.nytimes.com/2023/05/31/health/fda-rsv-vaccine-older-adults.html (accessed 2023-07-10).

(17) RSV (Respiratory Syncytial Virus) Immunizations | CDC. Centers for Disease Control and Prevention. https://www.cdc.gov/vaccines/vpd/rsv/index.html (accessed 2023-12-26).

(18) Miller, S. G.; Edwards, E. As RSV Cases Tick up, CDC Warns That a Key Drug to Keep Babies Safe Is in Short Supply. NBC News. October 23, 2023. https://www.nbcnews.com/health/health-news/rsv-cases-tick-key-drug-keep-babies-safe-short-supply-cdc-warns-rcna121763 (accessed 2023-12-26).

(19) Park, A. What to Know About the RSV Treatment Shortage. TIME. November 2, 2023. https://time.com/6330543/rsv-treatment-shortage/ (accessed 2023-12-26).

(20) RSV Surveillance Data - NREVSS | CDC. https://www.cdc.gov/surveillance/nrevss/rsv/index.html (accessed 2024-02-14).

(21) Borchers, A. T.; Chang, C.; Gershwin, M. E.; Gershwin, L. J. Respiratory Syncytial Virus— A Comprehensive Review. Clin. Rev. Allergy Immunol. 2013, 45 (3), 331–379. 10.1007/s12016-013-8368-9.

(22) Yu, J.; Liu, C.; Xiao, Y.; Xiang, Z.; Zhou, H.; Chen, L.; Shen, K.; Xie, Z.; Ren, L.; Wang, J. Respiratory Syncytial Virus Seasonality, Beijing, China, 2007–2015. Emerg. Infect. Dis. 2019, 25 (6), 1127–1135. 10.3201/eid2506.180532.

(23) Staadegaard, L.; Meijer, A.; Rodrigues, A. P.; Huang, S.; Cohen, C.; Demont, C.; van Summeren, J.; Caini, S.; Paget, J. Temporal Variations in Respiratory Syncytial Virus Epidemics, by Virus Subtype, 4 Countries. Emerg. Infect. Dis. 2021, 27 (5), 1537–1540. 10.3201/eid2705.204615.

(24) Graham, K. E.; Loeb, S. K.; Wolfe, M. K.; Catoe, D.; Sinnott-Armstrong, N.; Kim, S.; Yamahara, K. M.; Sassoubre, L. M.; Mendoza Grijalva, L. M.; Roldan-Hernandez, L.; Langenfeld, K.; Wigginton, K. R.; Boehm, A. B. SARS-CoV-2 RNA in Wastewater Settled Solids Is Associated with COVID-19 Cases in a Large Urban Sewershed. Environ. Sci. Technol. 2021, 55 (1), 488–498. 10.1021/acs.est.0c06191.

(25) Boehm, A. B.; Hughes, B.; Duong, D.; Chan-Herur, V.; Buchman, A.; Wolfe, M. K.; White, B. J. Wastewater Concentrations of Human Influenza, Metapneumovirus, Parainfluenza, Respiratory Syncytial Virus, Rhinovirus, and Seasonal Coronavirus Nucleic-Acids during the COVID-19 Pandemic: A Surveillance Study. Lancet Microbe 2023, 4 (5), e340–e348. 10.1016/S2666-5247(22)00386-X.

(26) Wolfe, M. K.; Duong, D.; Bakker, K. M.; Ammerman, M.; Mortenson, L.; Hughes, B.; Arts, P.; Lauring, A. S.; Fitzsimmons, W. J.; Bendall, E.; Hwang, C. E.; Martin, E. T.; White, B. J.; Boehm, A. B.; Wigginton, K. R. Wastewater-Based Detection of Two Influenza Outbreaks. Environ. Sci. Technol. Lett. 2022, 9 (8), 687–692. 10.1021/acs.estlett.2c00350.

(27) Wolfe, M. K.; Yu, A. T.; Duong, D.; Rane, M. S.; Hughes, B.; Chan-Herur, V.; Donnelly, M.; Chai, S.; White, B. J.; Vugia, D. J.; Boehm, A. B. Use of Wastewater for Mpox Outbreak Surveillance in California. N. Engl. J. Med. 2023, 388 (6), 570–572. 10.1056/NEJMc2213882.

(28) Hughes, B.; Duong, D.; White, B. J.; Wigginton, K. R.; Chan, E. M. G.; Wolfe, M. K.; Boehm, A. B. Respiratory Syncytial Virus (RSV) RNA in Wastewater Settled Solids Reflects RSV Clinical Positivity Rates. Environ. Sci. Technol. Lett. 2022, 9 (2), 173–178. 10.1021/acs.estlett.1c00963.

(29) Toribio-Avedillo, D.; Gómez-Gómez, C.; Sala-Comorera, L.; Rodríguez-Rubio, L.; Carcereny, A.; García-Pedemonte, D.; Pintó, R. M.; Guix, S.; Galofré, B.; Bosch, A.; Merino, S.; Muniesa, M. Monitoring Influenza and Respiratory Syncytial Virus in Wastewater. Beyond COVID-19. Sci. Total Environ. 2023, 892, 164495. 10.1016/j.scitotenv.2023.164495.

(30) Ahmed, W.; Bivins, A.; Stephens, M.; Metcalfe, S.; Smith, W. J. M.; Sirikanchana, K.; Kitajima, M.; Simpson, S. L. Occurrence of Multiple Respiratory Viruses in Wastewater in Queensland, Australia: Potential for Community Disease Surveillance. Sci. Total Environ. 2023, 864, 161023. 10.1016/j.scitotenv.2022.161023.

(31) Hayes, E. K.; Gouthro, M. T.; LeBlanc, J. J.; Gagnon, G. A. Simultaneous Detection of SARS-CoV-2, Influenza A, Respiratory Syncytial Virus, and Measles in Wastewater by Multiplex RT-qPCR. Sci. Total Environ. 2023, 889, 164261. 10.1016/j.scitotenv.2023.164261.

(32) Allen, D. M.; Reyne, M. I.; Allingham, P.; Levickas, A.; Bell, S. H.; Lock, J.; Coey, J. D.; Carson, S.; Lee, A. J.; McSparron, C.; Nejad, B. F.; McKenna, J.; Shannon, M.; Li, K.; Curran, T.; Broadbent, L. J.; Downey, D. G.; Power, U. F.; Groves, H. E.; McKinley, J. M.; McGrath, J. W.; Bamford, C. G. G.; Gilpin, D. F. Genomic Analysis and Surveillance of Respiratory Syncytial Virus (RSV) Using Wastewater-Based Epidemiology (WBE). medRxiv July 24, 2023, p 2023.07.21.23293016. 10.1101/2023.07.21.23293016.

(33) Koureas, M.; Mellou, K.; Vontas, A.; Kyritsi, M.; Panagoulias, I.; Koutsolioutsou, A.; Mouchtouri, V. A.; Speletas, M.; Paraskevis, D.; Hadjichristodoulou, C. Wastewater Levels of Respiratory Syncytial Virus Associated with Influenza-like Illness Rates in Children—A Case Study in Larissa, Greece (October 2022–January 2023). Int. J. Environ. Res. Public. Health 2023, 20 (6), 5219. 10.3390/ijerph20065219.

(34) Rector, A.; Bloemen, M.; Thijssen, M.; Pussig, B.; Beuselinck, K.; Van Ranst, M.; Wollants, E. Respiratory Viruses in Wastewater Compared with Clinical Samples, Leuven, Belgium. Emerg. Infect. Dis. 2024, 30 (1), 141–145. 10.3201/eid3001.231011.

(35) Mercier, E.; Pisharody, L.; Guy, F.; Wan, S.; Hegazy, N.; D’Aoust, P. M.; Kabir, M. P.; Nguyen, T. B.; Eid, W.; Harvey, B.; Rodenburg, E.; Rutherford, C.; Mackenzie, A. E.; Willmore, J.; Hui, C.; Paes, B.; Delatolla, R.; Thampi, N. Wastewater-Based Surveillance Identifies Start to the Pediatric Respiratory Syncytial Virus Season in Two Cities in Ontario, Canada. Front. Public Health 2023, 11, 1261165. 10.3389/fpubh.2023.1261165.

(36) Robertson, M.; Eden, J.-S.; Levy, A.; Carter, I.; Tulloch, R. L.; Cutmore, E. J.; Horsburgh, B. A.; Sikazwe, C. T.; Dwyer, D. E.; Smith, D. W.; Kok, J. The Spatial-Temporal Dynamics of Respiratory Syncytial Virus Infections across the East–West Coasts of Australia during 2016–17. Virus Evol. 2021, 7 (2), veab068. 10.1093/ve/veab068.

(37) Agoti, C. N.; Otieno, J. R.; Ngama, M.; Mwihuri, A. G.; Medley, G. F.; Cane, P. A.; Nokes, D. J. Successive Respiratory Syncytial Virus Epidemics in Local Populations Arise from Multiple Variant Introductions, Providing Insights into Virus Persistence. J. Virol. 2015, 89 (22), 11630–11642. 10.1128/jvi.01972-15.

(38) Rozenbaum, M. H.; Begier, E.; Kurosky, S. K.; Whelan, J.; Bem, D.; Pouwels, K. B.; Postma, M.; Bont, L. Incidence of Respiratory Syncytial Virus Infection in Older Adults: Limitations of Current Data. Infect. Dis. Ther. 2023, 12 (6), 1487–1504. 10.1007/s40121-023-00802-4.

(39) Zulli, A.; Varkila, M. R. J.; Parsonnet, J.; Wolfe, M. K.; Boehm, A. B. Observations of Respiratory Syncytial Virus (RSV) Nucleic Acids in Wastewater Solids Across the United States in the 2022–2023 Season: Relationships with RSV Infection Positivity and Hospitalization Rates. ACS EST Water 2024. 10.1021/acsestwater.3c00725.

(40) Lowry, S. A.; Wolfe, M. K.; Boehm, A. B. Respiratory Virus Concentrations in Human Excretions That Contribute to Wastewater: A Systematic Review and Meta-Analysis. J. Water Health 2023, 21 (6), 831–848. 10.2166/wh.2023.057.

(41) Bureau, U. C. Large Southern Cities Lead Nation in Population Growth. Census.gov. https://www.census.gov/newsroom/press-releases/2023/subcounty-metro-micro-estimates.html (accessed 2024-01-16).

(42) Esri; TomTom; Garmin; FAO; NOAA; USGS; © OpenStreetMap contributors; GIS User Community. Light Gray Canvas Base, 2017. https://basemaps.arcgis.com/arcgis/rest/services/World_Basemap_v2/VectorTileServer (accessed 2024-03-05).

(43) Walters, S. J.; Campbell, M. J. The Use of Bootstrap Methods for Analysing Health-Related Quality of Life Outcomes (Particularly the SF-36). Health Qual. Life Outcomes 2004, 2, 70. 10.1186/1477-7525-2-70.

(44) Zambrana, W.; Huang, C.; Solis, D.; Sahoo, M. K.; Pinsky, B. A.; Boehm, A. B. Data on Spatial and Temporal Variation in Respiratory Syncytial Virus (RSV) Subtype RNA in Wastewater, and Relation to Clinical Specimens. 2024. 10.25740/jb062yw6825.

(45) Huisman, J. S.; Scire, J.; Caduff, L.; Fernandez, -Cassi Xavier; Ganesanandamoorthy, P.; Kull, A.; Scheidegger, A.; Stachler, E.; Boehm, A. B.; Hughes, B.; Knudson, A.; Topol, A.; Wigginton, K. R.; Wolfe, M. K.; Kohn, T.; Ort, C.; Stadler, T.; Julian, T. R. Wastewater-Based Estimation of the Effective Reproductive Number of SARS-CoV-2. Environ. Health Perspect. 2022, 130 (5), 057011. 10.1289/EHP10050.

(46) Wolfe, M. K.; Topol, A.; Knudson, A.; Simpson, A.; White, B.; Vugia, D. J.; Yu, A. T.; Li, L.; Balliet, M.; Stoddard, P.; Han, G. S.; Wigginton, K. R.; Boehm, A. B. High-Frequency, High-Throughput Quantification of SARS-CoV-2 RNA in Wastewater Settled Solids at Eight Publicly Owned Treatment Works in Northern California Shows Strong Association with COVID-19 Incidence. mSystems 2021, 0 (0), e00829–21. 10.1128/mSystems.00829-21.

(47) Topol, A.; Wolfe, M.; White, B.; Wigginton, K.; Boehm, A. B. High Throughput pre-analytical processing of wastewater settled solids for SARS-CoV-2 RNA analyses. protocols.io. https://www.protocols.io/view/high-throughput-pre-analytical-processing-of-waste-b2kmqcu6 (accessed 2022-04-15).

(48) Decaro, N.; Elia, G.; Campolo, M.; Desario, C.; Mari, V.; Radogna, A.; Colaianni, M. L.; Cirone, F.; Tempesta, M.; Buonavoglia, C. Detection of Bovine Coronavirus Using a TaqMan-Based Real-Time RT-PCR Assay. J. Virol. Methods 2008, 151 (2), 167–171. 10.1016/j.jviromet.2008.05.016.

(49) Zhang, T.; Breitbart, M.; Lee, W. H.; Run, J.-Q.; Wei, C. L.; Soh, S. W. L.; Hibberd, M. L.; Liu, E. T.; Rohwer, F.; Ruan, Y. RNA Viral Community in Human Feces: Prevalence of Plant Pathogenic Viruses. PLOS Biol. 2005, 4 (1), e3. 10.1371/journal.pbio.0040003.

(50) Haramoto, E.; Kitajima, M.; Kishida, N.; Konno, Y.; Katayama, H.; Asami, M.; Akiba, M. Occurrence of Pepper Mild Mottle Virus in Drinking Water Sources in Japan. Appl. Environ. Microbiol. 2013, 79 (23), 7413–7418. 10.1128/AEM.02354-13.

(51) Boehm, A. B.; Wolfe, M. K.; White, B. J.; Hughes, B.; Duong, D.; Banaei, N.; Bidwell, A. Human Norovirus (HuNoV) GII RNA in Wastewater Solids at 145 United States Wastewater Treatment Plants: Comparison to Positivity Rates of Clinical Specimens and Modeled Estimates of HuNoV GII Shedders. J. Expo. Sci. Environ. Epidemiol. 2023. 10.1038/s41370-023-00592-4.

(52) Topol, A.; Wolfe, M.; Wigginton, K.; White, B.; Boehm, A. High Throughput RNA Extraction and PCR Inhibitor Removal of Settled Solids for Wastewater Surveillance of S… 2021.

(53) Topol, A.; Wolfe, M.; White, B.; Wigginton, K.; Boehm, A. B. High Throughput SARS-COV-2, PMMOV, and BCoV Quantification in Settled Solids Using Digital RT-PCR. 2022.

(54) Wang, L.; Piedra, P. A.; Avadhanula, V.; Durigon, E. L.; Machablishvili, A.; López, M.-R.; Thornburg, N. J.; Peret, T. C. T. Duplex Real-Time RT-PCR Assay for Detection and Subgroup-Specific Identification of Human Respiratory Syncytial Virus. J. Virol. Methods 2019, 271, 113676. 10.1016/j.jviromet.2019.113676.

(55) Borchardt, M. A.; Boehm, A. B.; Salit, M.; Spencer, S. K.; Wigginton, K. R.; Noble, R. T. The Environmental Microbiology Minimum Information (EMMI) Guidelines: qPCR and dPCR Quality and Reporting for Environmental Microbiology. Environ. Sci. Technol. 2021, 55 (15), 10210–10223. 10.1021/acs.est.1c01767.

(56) Reese, P. E.; Marchette, N. J. Respiratory Syncytial Virus Infection and Prevalence of Subgroups A and B in Hawaii. J. Clin. Microbiol. 1991, 29 (11), 2614–2615. 10.1128/jcm.29.11.2614-2615.1991.

(57) Pitzer, V. E.; Viboud, C.; Alonso, W. J.; Wilcox, T.; Metcalf, C. J.; Steiner, C. A.; Haynes, A. K.; Grenfell, B. T. Environmental Drivers of the Spatiotemporal Dynamics of Respiratory Syncytial Virus in the United States. PLOS Pathog. 2015, 11 (1), e1004591. 10.1371/journal.ppat.1004591.

(58) Zheng, Z.; Pitzer, V. E.; Warren, J. L.; Weinberger, D. M. Community Factors Associated with Local Epidemic Timing of Respiratory Syncytial Virus: A Spatiotemporal Modeling Study. Sci. Adv. 2021, 7 (26), eabd6421. 10.1126/sciadv.abd6421.

(59) Guo, Z.; Xiao, G.; Wang, Y.; Li, S.; Du, J.; Dai, B.; Gong, L.; Xiao, D. Dynamic Model of Respiratory Infectious Disease Transmission in Urban Public Transportation Systems. Heliyon 2023, 9 (3), e14500. 10.1016/j.heliyon.2023.e14500.

(60) AHPS Precipitation Analysis. https://water.weather.gov/precip/ (accessed 2024-01-08).

(61) National Maps | National Centers for Environmental Information (NCEI). National Oceanic and Atmospheric Administration. https://www.ncei.noaa.gov/access/monitoring/us-maps/ (accessed 2024-01-08).

(62) U.S. Census Bureau. Census Bureau Data. https://data.census.gov/ (accessed 2024-01-08).

(63) U.S. Department of Transportation. Federal Highway Administration. Status of the Nation’s Highways, Bridges, and Transit: Conditions and Performance, 24th Edition. 10.21949/1521794.

(64) Hamid, S.; Winn, A.; Parikh, R.; Jones, J. M.; McMorrow, M.; Prill, M. M.; Silk, B. J.; Scobie, H. M.; Hall, A. J. Seasonality of Respiratory Syncytial Virus — United States, 2017–2023. MMWR Morb. Mortal. Wkly. Rep. 2023, 72 (14), 355–361. 10.15585/mmwr.mm7214a1.

