## Supporting material for "Spatial and temporal variation in respiratory syncytial virus (RSV) subtype RNA in wastewater, and relation to clinical specimens"

#### 1. Materials and Methods

##### Study Design

The samples in this study are part of a larger COVID-19 wastewater surveillance effort which, alongside several viruses, measures total respiratory syncytial virus (RSV) (sum of RSV A and RSV B, not subtyped) routinely[1]. Total RSV was measured in fresh samples alongside PMMoV and BCoV, as described in the main text. Sequences of primers and probes for Total RSV used in the larger epidemiology effort can be found in **Table S1**. Total RSV results are not reported in this study, and they were only used for original sample selection and storage impact assessment.

##### Procedures

The primers and probes sequences used in the study and their corresponding references can be found in **Table S1-S2**.

#### 2. Results

Concentrations of RSV A and RSV B at each wastewater treatment plant are shown in **Figure S1**, and the proportion of RSV A to Total RSV at each wastewater treatment plant is shown in **Figure S2**.

## QA/QC.

The Environmental Microbiology Minimal Information (EMMI) guidelines checklist can be found in **Figure S3**, and examples of fluorescent plots for this study can be found in **Figure S4**.

#### 3. References


(1) Zulli, A.; Varkila, M. R. J.; Parsonnet, J.; Wolfe, M. K.; Boehm, A. B. Observations of Respiratory Syncytial Virus (RSV) Nucleic Acids in Wastewater Solids Across the United States in the 2022–2023 Season: Relationships with RSV Infection Positivity and Hospitalization Rates. *ACS EST Water* **2024**. https://doi.org/10.1021/acsestwater.3c00725.

(2) Borchardt, M. A.; Boehm, A. B.; Salit, M.; Spencer, S. K.; Wigginton, K. R.; Noble, R. T. The Environmental Microbiology Minimum Information (EMMI) Guidelines: qPCR and dPCR Quality and Reporting for Environmental Microbiology. *Environ. Sci. Technol.* **2021**, *55* (15), 10210–10223. https://doi.org/10.1021/acs.est.1c01767.

(3) Borchardt, M.; Boehm, A.; Wigginton, K.; Noble, R.; Salit, M.; Spencer, S. Environmental Microbiology Minimal Information Checklist. **2023**. https://doi.org/10.25740/TM549RG8921.

(4) Boehm, A. B.; Hughes, B.; Duong, D.; Chan-Herur, V.; Buchman, A.; Wolfe, M. K.; White, B. J. Wastewater Concentrations of Human Influenza, Metapneumovirus, Parainfluenza, Respiratory Syncytial Virus, Rhinovirus, and Seasonal Coronavirus Nucleic-Acids during the COVID-19 Pandemic: A Surveillance Study. *Lancet Microbe* **2023**, *4* (5), e340–e348. https://doi.org/10.1016/S2666-5247(22)00386-X.

(5) Haramoto, E.; Kitajima, M.; Kishida, N.; Konno, Y.; Katayama, H.; Asami, M.; Akiba, M. Occurrence of Pepper Mild Mottle Virus in Drinking Water Sources in Japan. *Appl. Environ. Microbiol.* **2013**, *79* (23), 7413–7418. https://doi.org/10.1128/AEM.02354-13.

(6) Zhang, T.; Breitbart, M.; Lee, W. H.; Run, J.-Q.; Wei, C. L.; Soh, S. W. L.; Hibberd, M. L.; Liu, E. T.; Rohwer, F.; Ruan, Y. RNA Viral Community in Human Feces: Prevalence of Plant Pathogenic Viruses. *PLOS Biol.* **2005**, *4* (1), e3. https://doi.org/10.1371/journal.pbio.0040003.

(7) Decaro, N.; Elia, G.; Campolo, M.; Desario, C.; Mari, V.; Radogna, A.; Colaianni, M. L.; Cirone, F.; Tempesta, M.; Buonavoglia, C. Detection of Bovine Coronavirus Using a TaqMan-Based Real-Time RT-PCR Assay. *J. Virol. Methods* **2008**, *151* (2), 167–171. https://doi.org/10.1016/j.jviromet.2008.05.016.

(8) Hughes, B.; Duong, D.; White, B. J.; Wigginton, K. R.; Chan, E. M. G.; Wolfe, M. K.; Boehm, A. B. Respiratory Syncytial Virus (RSV) RNA in Wastewater Settled Solids Reflects RSV Clinical Positivity Rates. *Environ. Sci. Technol. Lett.* **2022**, *9* (2), 173–178. https://doi.org/10.1021/acs.estlett.1c00963.

### Figures

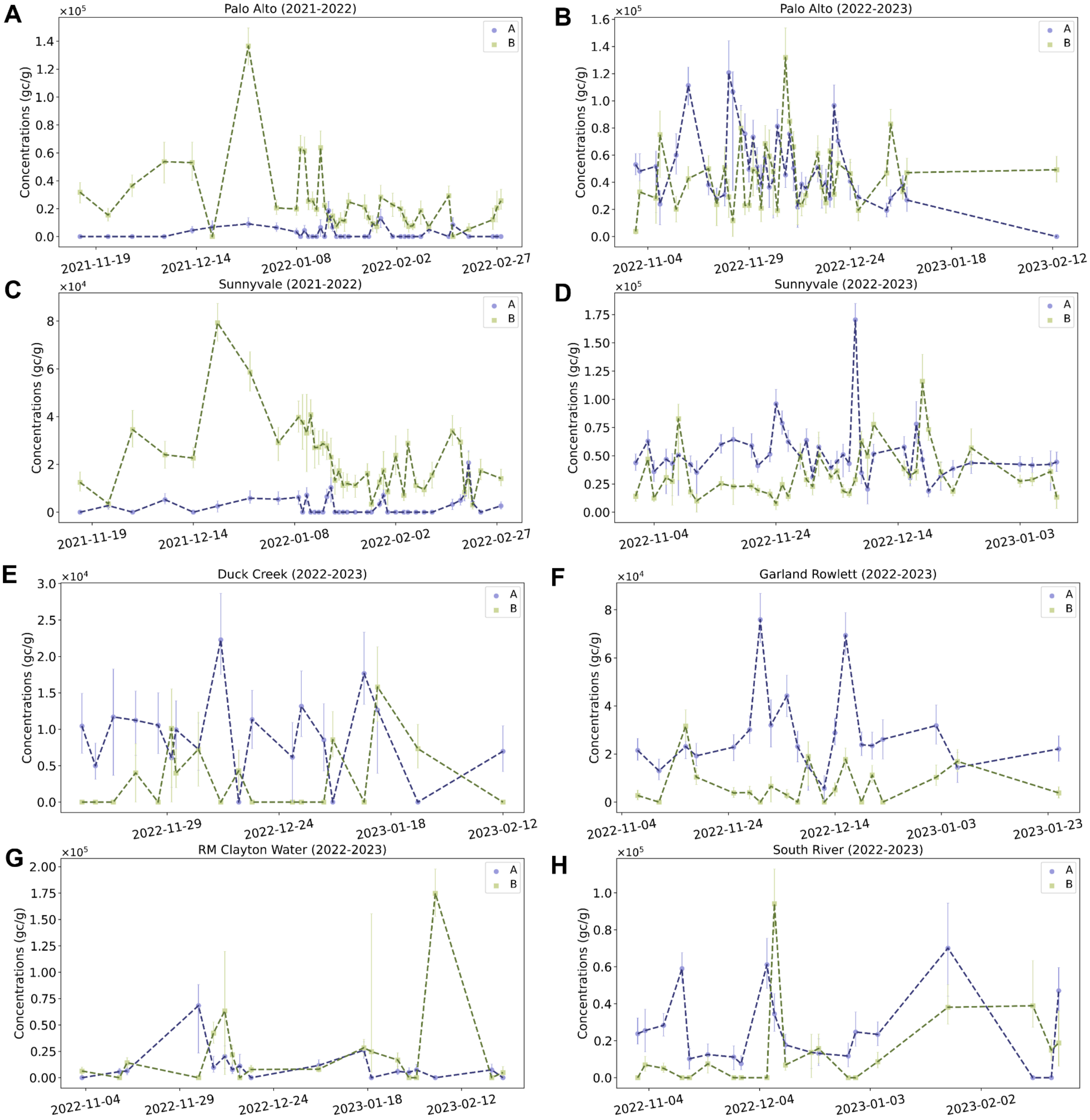


**Figure S1**. RSV A (purple) and RSV B (green) concentrations for all WWTPs included in the study. Palo Alto Season 1 [San Jose, CA] (A) ; Palo Alto Season 2 [San Jose, CA] (B); Sunnyvale Season 1 [San Jose, CA] (C); Sunnyvale Season 2 [San Jose, CA] (D); Duck Creek Season 2 [Dallas, TX] (E); Garland Rowlett Season 2 [Dallas,

TX] (F); RM Clayton Water Season 2 [Atlanta, GA] (G); South River Season 2 [Atlanta, GA] (H). Error bars represent the standard deviation as the total error reported by the ddPCR instrument.

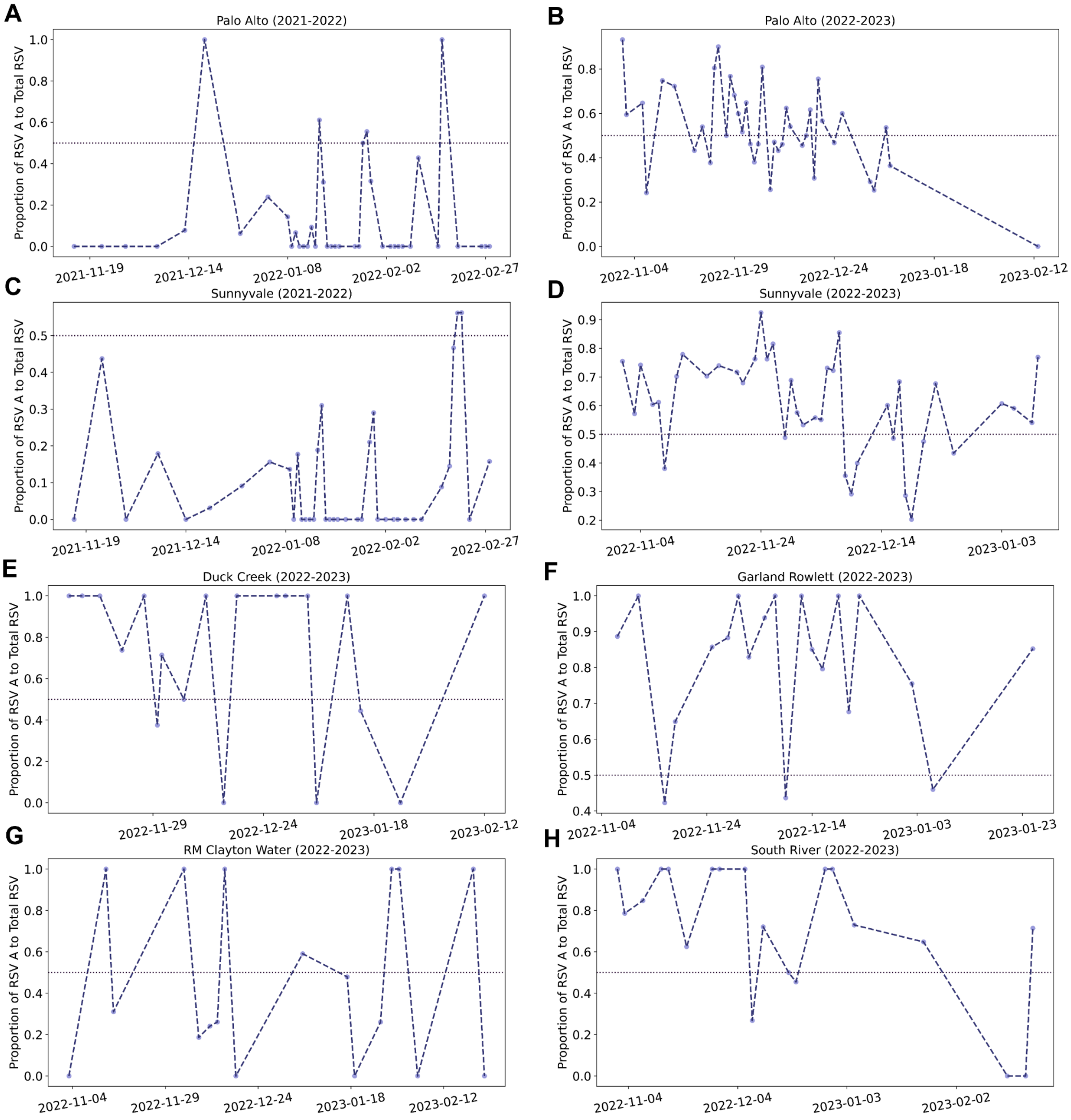


**Figure S2**. Proportion of RSV A to total RSV for all WWTPs included in the study. Palo Alto Season 1 [San Jose, CA] (A) ; Palo Alto Season 2 [San Jose, CA] (B); Sunnyvale Season 1 [San Jose, CA] (C); Sunnyvale Season 2 [San

Jose, CA] (D); Duck Creek Season 2 [Dallas, TX] (E); Garland Rowlett Season 2 [Dallas, TX] (F); RM Clayton Water Season 2 [Atlanta, GA] (G); South River Season 2 [Atlanta, GA] (H). Dashed horizontal line indicates a proportion of 0.5.

| Study Description | Environmental Sampling | Sample Treatment | Sample Reduction | Nucleic-acid Extraction | Reverse Transcription | PCR Amplification | Analysis |
|---|---|---|---|---|---|---|---|
| Study name: RSV A/B in w<br>Date: January 10-2024<br>Completed by: Winnie Zam | Notes: As described in text | Notes: Performed for wastewater solids samples - Samples were frozen and stored as described in the text. Information on the effect on samples through total RSV and PMMoV concentrations provided. | Notes: Performed for wastewater solids samples - Centrifugation and resuspension as described n text | Notes: As described in text | Notes: Performed -As described in text | Notes: As described in text. No specific control just for the PCR step. | Notes: As described in text. |

| Control Checklist | Environmental Sampling | Sample Treatment | Sample Reduction | Nucleic-acid Extraction | Reverse Transcription | PCR Amplification | |
|---|---|---|---|---|---|---|---|
| Step performed | ☑ | ☑ | ☑ | ☑ | ☑ | ☑ | |
| Step has control info | ☐ | ☐ | ☐ | ☑ | ☑ | ☐ | Negative controls |
| # of control replicates | NA | NA | NA | 2 | 2 | NA | |
| Control result reported | ☐ | ☐ | ☐ | ☑ | ☑ | ☐ | |
| Method for handling failed controls described | ☐ | ☐ | ☐ | ☑ | ☑ | ☐ | |
| Step has control info | ☐ | ☐ | ☐ | ☐ | ☐ | ☐ | Positive controls |
| Control identity described | ☐ | ☐ | ☐ | ☑ | ☑ | ☐ | |
| Control quantification method described | ☐ | ☑ | ☐ | ☑ | ☑ | ☐ | |
| # control replicates | NA | NA | NA | 10 | 2 | NA | |
| Control result reported | ☐ | ☑ | ☐ | ☑ | ☑ | ☐ | |
| Method for handling failed controls described | ☐ | ☐ | ☐ | ☑ | ☑ | ☐ | |

Process checklist

| Environmental Sampling | |
|---|---|
| Sample procedure | ☑ |
| Number of samples | ☑ |
| Sample amount, mean, range | ☑ |
| Sampling locations, dates, times | ☑ |
| Sample storage conditions | ☑ |

| Sample Treatment | |
|---|---|
| Treatment procedure | ☑ |
| Reagents | ☐ |

| Sample Reduction | |
|---|---|
| Reduction procedure | ☑ |
| Reagents | ☐ |
| Concentration factor | ☐ |

| Nucleic acid Extraction | |
|---|---|
| Extraction procedure | ☑ |
| Volume or mass extracted, volume or mass obtained | ☐ |
| Extract storage conditions | ☑ |

| Reverse Transcription | |
|---|---|
| One- or two-step | ☑ |
| cDNA storage conditions (if 2 step) | ☐ |
| Reaction temperatures and times | ☑ |
| Reaction reagents and concentrations | ☑ |
| Priming method | ☑ |
| Reaction volume, added template amount | ☑ |
| RT efficiency assessment procedure (if 2-step) | ☐ |
| RT control description (if two-step) | ☐ |
| RT efficiency reported (if 2-step) | ☐ |

| qPCR or dPCR | |
|---|---|
| Target gene name, amplicon length | ☑ |
| Thermocycling temp and times | ☑ |
| Master mix composition: vendors, concentrations | ☑ |
| Additives: vendors, composition | ☑ |
| Template amount added, pre-treatment (if any) | ☑ |
| Primers: sequences, concentrations, vendors, references | ☑ |
| Amplicon confirmation method (probe, melt curve details, etc) | ☐ |
| Probe sequence, concentration, vendor, reference | ☑ |
| Instrumentation | ☑ |
| Inhibition assessment procedure | ☑ |
| Inhibition control description (if used) | ☐ |
| Number of samples tested and found inhibited | ☐ |
| Equivalent volume of sample analyzed | ☐ |

| Analysis- dPCR | |
|---|---|
| Threshold settings | ☑ |
| Technical replicates, number, well merging | ☑ |
| Partitions measured, number, mean, variance | ☑ |
| Partition volume | ☑ |
| Target copies per partition, mean, variance | ☑ |
| Program used for dPCR analysis | ☑ |
| Explanation of control results, example plots | ☑ |

| Analysis- qPCR | |
|---|---|
| Technical replicates, number, calculations | ☑ |
| Calibration standards, description, source | ☑ |
| Method of quantifying standards | ☑ |
| Calibration curve slope | ☐ |
| Calibration curve R2 | ☐ |
| Lowest standard measured or 95% LOD | ☐ |
| Cq value determination methods | ☑ |

Note to users: This checklist is provided as guidance for best practices for reporting, but is not meant to be prescriptive. Not all items in the check list will apply to all studies. Please see Borchardt et al. The Environmental Microbiology Minimum Information (EMMI) Guidelines: qPCR and dPCR Quality and Reporting for Environmental Microbiology. Environmental Science & Technology, 2021, 55, 15, 10210–10223.

Version 3.0

This version maintained by Borchardt, Boehm, Salit, Noble, Wiggington, Spencer

Date: 18 October 2023

**Figure S3**. Environmental Microbiology Minimal Information (EMMI) guidelines checklist[2,3].

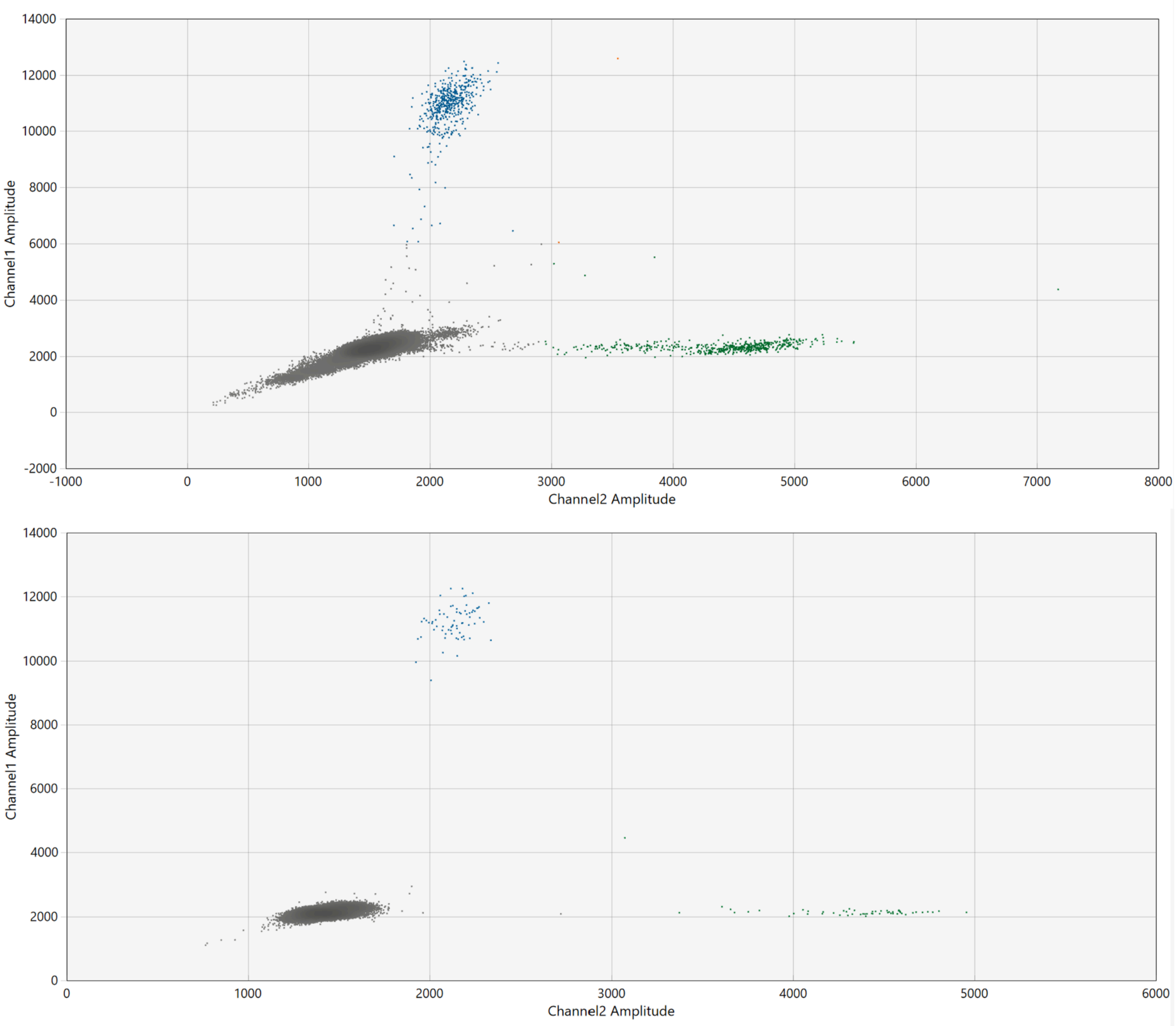


**Figure S4**. Examples of ddPCR experimental results for the RSV A/B assay. Several wastewater representative samples (top) and positive controls only (bottom).

#### Tables

**Table S1**. Primer and probe sequences used in this study for wastewater samples

| Target | Forward Primer Sequence | Reverse Primer Sequence | Probe Sequence | Reference |
|---|---|---|---|---|
| RSV A | AGAGGTGGCAGTAGAGTTGA | CTCCACAACTTGTTCCATTTCTG | ATGGTGCAGGGCAAGTGATG (5’ HEX/ZEN/3’ IBFQ) | Boehm et al[4] |
| RSV B | TGACACTCCCAATTATGATGTGC | CCTGTGAATTTATGATTTGCATCTTCAG | ACACCTAAACAAACTATGTGGTATGC (5’ FAM/ZEN/3’ IBFQ) | Boehm et al[4] |
| PMMoV[a,b] | GAGTGGTTTGACCTTAACGTTTGA | TTGTCGGTTGCAATGCAAGT | CCTACCGAAGCAAATG (5’ HEX/ZEN/3’ IBFQ) | Haramoto et al[5] Zhang et al[6] |
| BCoV[a] | CTGGAAGTTGGTGGAGTT | ATTATCGGCCTAACATACATC | CCTTCATATCTATACACATCAAGTTGTT (5’ FAM/ZEN/3’ IBFQ) | Decaro et al[7] |
| Total RSV[a,c] | TCCAGAATAYAGGC-ATGAYTCTCC | CYCTYCTAATYACWGCTGTAAGAC | AACCAAATTAGCAGCAGGA-GATAGATCAG (5′HEX/ZEN/3′IBFQ) | Hughes et al[8] |

Note: a = Measured as part of the larger epidemiology effort; b = Additional PMMoV measurements were conducted in singleplex for a subset of samples concurrently with RSV A/B measurements to assess the impact of freeze-thaw cycles; c = Only used in this study to assess the impact of freeze-thaw cycles. The sum of RSV A and RSV B measurements were compared against total RSV measurements with no storage or freeze-thaw cycles.

**Table S2**. Primer and probes sequences used in this study for clinical samples

| Name | Sequence (5'to 3') | Final Concentration |
|---|---|---|
| RSV-F Primer | ATGGCTCTTAGCAAAGTCAAGT | 400 nM |
| RSV-R Primer | TGCACATCATAATTRGGAGTRTCA | 400 nM |
| RSV A-Probe | FAM-ACACTCAAC /ZEN/AAAGAT CAACTTCTRTCATCCAGCA-3IABkFQ | 200 nM |
| RSV B-Probe | Cy5-ACATTAAAT /TAO/AAGGATCAG CTGCTGTCATCCAGCA-3IAbRQSp | 200 nM |
| RNaseP Forward Primer | AGATTTGGACCTGCGAGCG | 100 nM |
| RNaseP Reverse Primer | GAGCGGCTGTCTCCACAAGT | 100 nM |
| RNaseP Probe | Cal Fluor 560-TTCTGACCTGAAGGCTCTGCGCG-BHQ-1 | 50 nM |

Note:BHQ, Black Hole Quencher; Cy5, Cyanine-5; FAM, 6-carboxyfluorescein; RSV, Respiratory Syncytial Virus. RSV primers and probes were purchased from Integrated DNA Technologies (IDT; San Diego, California). The probes contain internal quenchers ZEN and TAO in addition to the 3' Iowa Black quenchers FQ (3IABkFQ) and RQ (3IAbRQSp) (all proprietary to IDT). RnaseP primers and probe were purchased from Biosearch Technologies (Petaluma, California).

**Table S3**. Overall $P_{A,WW}$ results and statistical analysis for the Kruskal-Wallis test

| Group | Number of samples (n) | Median $P_{A,WW}$ | $P_{A,WW}$ IQR | Kruskal-Wallis P-Value |
|---|---|---|---|---|
| **Spatial (n = 157)** | | | | |
| Santa Clara County, CA[a] | 80 | 0.58 | 0.26 | $1.19 \times 10^{-4}$* |
| Dallas, TX | 39 | 0.88 | 0.34 | |
| Atlanta, GA | 38 | 0.68 | 0.74 | |
| **Temporal (n = 160)** | | | | |
| Sunnyvale WWTP (Season 1) | 40 | 0.00 | 0.16 | $1.47 \times 10^{-19}$* |
| Palo Alto WWTP (Season 1) | 40 | 0.00 | 0.11 | |
| Sunnyvale WWTP (Season 2) | 40 | 0.61 | 0.21 | |
| Palo Alto WWTP (Season 2) | 40 | 0.53 | 0.21 | |
| **Clinical (n = 169)** | | | | |
| Wastewater solid samples[a] | 80 | 0.58 | 0.26 | $3.49 \times 10^{-19}$* |
| Clinical samples bootstrap[b] | 80 | 0.79 | 0.03 | |

Note: $P_{A,WW}$ = proportion of RSV A to total RSV in wastewater solids; IQR =interquartile range ;* = Statistically significant per the Kruskal-Wallis test with a significance level of p =0.006 accounting for the Bonferroni correction; a = The samples included in the groups 'Santa Clara County, CA' and 'Wastewater samples' are the same samples for Sunnyvale WWTP Season 2 and Palo Alto WWTP Season 2 used in the Temporal evaluation; b = The numbers reported on this table were obtained from bootstrapping method (median bootstrap $F_{A,Cl}$ and $F_{A,Cl}$ IQR). The total number of clinical samples tested was 593 with 79% of samples testing positive for RSV A.

**Table S4**. Statistical analysis results for the Conover-Iman test

| ***Spatial (n = 157)*** | **Atlanta, GA** | **Dallas, TX** | |
|---|---|---|---|
| **Dallas, TX** | (-2.57)<br>$5.49 \times 10^{-3}$* | - | |
| **Santa Clara County, CA**[a] | (1.47)<br>$7.13 \times 10^{-2}$ | (4.49)<br>$6.89 \times 10^{-6}$* | |
| ***Temporal (n = 160)*** | **Palo Alto (Season 1)** | **Palo Alto (Season 2)** | **Sunnyvale (Season 1)** |
| **Palo Alto (Season 2)** | (-8.95)<br>$5.06 \times 10^{-16}$* | - | - |
| **Sunnyvale (Season 1)** | (0.35)<br>$3.62 \times 10^{-1}$ | (9.30)<br>$6.01 \times 10^{-17}$* | - |
| **Sunnyvale (Season 2)** | (-10.89)<br>$3.36 \times 10^{-21}$* | (-1.94)<br>$2.68 \times 10^{-2}$ | (-11.24)<br>$3.68 \times 10^{-22}$* |
| ***Clinical (n = 160)*** | **Clinical Samples**[b] | | |
| **Wastewater Samples**[a] | (12.67)<br>$3.88 \times 10^{-26}$* | | |

Note: P-values for the Conover-Iman test with corresponding test statistics in parentheses. The sign of the test statistic is used to indicate the directionality of the differences where a positive test statistic value indicates Column Category> Row Category, negative test statistic value indicates Column Category < Row Category; * = Statistically significant per the Conover-Iman test with a significance level of p =0.006 accounting for the Bonferroni correction; a = The samples included in the groups 'Santa Clara County, CA' and 'Wastewater samples' are the same samples for Sunnyvale WWTP Season 2 and Palo Alto WWTP Season 2 used in the Temporal evaluation; b = The numbers reported on this table were obtained from bootstrapping method (using median bootstrap $F_{A, Cl}$). The total number of clinical samples tested was 593 with 79% of samples testing positive for RSV A.
